# “ARVs is for HIV and cream is for HPV or precancer:” Women’s Perceptions and Perceived Acceptability of Self-Administered Topical Therapies for Cervical Precancer Treatment: A Qualitative Study from Kenya

**DOI:** 10.1101/2024.03.11.24304083

**Authors:** Chemtai Mungo, Aparna Ghosh Kachoria, Everlyn Adoyo, Graham Zulu, Supreet Kaur Goraya, Jackton Omoto, Cirilus Osongo, Renée M. Ferrari, Lisa Rahangdale

## Abstract

**Background:** Women in low- and middle-income countries (LMICs) bear a disproportionate burden of cervical cancer, despite being a preventable disease. Prevention efforts in LMICs are hindered in part by lack of access to cervical precancer treatment, due to weak health infrastructure and a lack of adequate human resources to deliver current provider-administered precancer treatments. Innovative strategies are urgently needed to close the cervical precancer treatment gap in LMICs, including the use of self-administered topical therapies such as 5-fluorouracil and imiquimod, for which efficacy evidence is available from high-income settings. We investigated African women’s perceptions and perceived acceptability of these therapies for cervical precancer treatment.

**Methods:** Between November 2022 and April 2023, we conducted five focus group discussions (FGDs) with women ages 25-65 years undergoing cervical cancer screening or precancer treatment in Kisumu, Kenya. The FGDs explored women’s experiences with screening and precancer treatment, their acceptability of topical therapies for precancer treatment, and perceived barriers and facilitators to uptake. The FGDs were moderated by local qualitative research assistants, conducted in local languages, transcribed, coded, and analyzed using qualitative description using NVIVO software.

**Results:** Twenty-nine women participated, with a mean age of 35.4 years (SD 6.5). All had undergone cervical cancer screening, and 25 (83%) had a history of precancer treatment with ablation or excision. Multiple themes were identified related to women’s perceptions of topical therapies. Participants were highly receptive of topical treatments, with many favoring the option of self-administration compared to provider-administration of such therapies. Self-administration of topical therapies was felt to help address challenges associated with current treatment methods, including difficulty in access, pain with procedures, cost, and lack of privacy with pelvic exams. Participants had a preference for topical therapies that are used less frequently compared to those used daily.

**Conclusions:** Among Kenyan women with a history of cervical precancer treatment, self-administered topical therapies for precancer are acceptable and have the potential to address barriers, including access, privacy, and cost, that hinder precancer treatment in LMICs. If supported by efficacy studies in LMICs, self-administered topical therapies offer a scalable approach to closing the precancer treatment gap in LMICs.

**Trial registration:** *Not applicable*

## Background

Women in low- and middle-income countries (LMICs) shoulder a disproportionate burden of the incidence and mortality from cervical cancer, accounting for 85 percent of cases and 90 percent of deaths in 2020[1]. Additionally, women living with HIV (WLWH), the majority of whom live in LMICs, are six times more likely to develop cervical cancer and, hence, are a priority population for prevention[2], [3]. In response to this, the World Health Organization (WHO) launched the 90/70/90 global strategy to eliminate cervical cancer[4]. This strategy, adopted by most WHO member states, calls for 90% human papillomavirus (HPV) vaccination coverage of all girls by the age 15 years, 70% of women globally receiving cervical cancer screening with a high-performance test at least twice in their lifetime, and 90% of those with a positive result adequately treated by 2030[4]. Modeling studies demonstrate that achieving the 90/70/90 targets will avert 74 million new cases of cervical cancer and 62 million deaths in LMICs alone[5].

Among unvaccinated women, cervical cancer can be prevented through screening for and treating early changes in the cervix, known as cervical precancer, caused by HPV infection. Current cervical precancer treatment options include ablation or excision procedures, both of which are performed by trained healthcare professionals[6]. Despite progress in screening, access to cervical precancer treatment following abnormal screening results in LMICs is highly limited[7], [8], [9], [10][11]. In a review of the Kenya national cervical cancer screening program in 2021, only 26% of 10,983 women who screened positive for cervical precancer received treatment [12]. Similarly, between 2011 and 2015 in Malawi, only 43.3% and 31.8% of women with cervical precancer who required ablation or excision, respectively, received treatment[13]. Challenges associated with precancer treatment in LMICs include high rates of loss-to-follow-up due to cost and transportation challenges when women screened in rural areas are referred to central facilities where treatment is available, due to a lack of skilled healthcare providers in rural areas where most women live [9], [10], [11], [12], [14], [15]. The failure to treat precancerous lesions while at a curable stage in these settings results in 85% of new global cervical cancer cases occurring in LMICs, highlighting a significant disparity. This highlights the urgent need for innovative yet resource-appropriate approaches to address the gap in cervical precancer treatment in LMICs. One potential strategy is the use of self-administered topical therapies.

While no topical therapies are currently approved for the treatment of cervical precancer, the use of self- or provider-administered topical therapies for cervical precancer treatment is an area of active investigation [16], [17], [18], [19], [20], [21], [22], [23]. The feasibility, acceptability, and efficacy of topical therapies for cervical precancer treatment has been demonstrated by several studies in high-income countries, including randomized trials[16], [17], [20], [24]. Several of these drugs are on the WHO List of Essential Medications and are readily available in LMICs in generic form[25]. One such drug is Fluorouracil (5FU) cream, which has been demonstrated to be a safe and effective cervical precancer treatment when self-administered intravaginally[16], [17]. Compared to provider-administered precancer treatment, which is currently inaccessible for many women in LMICs, patient-administered therapies may be a highly scalable and cost-effective cervical precancer treatment method in these settings.

To inform ongoing (Clinicaltrials.gov identifier NCT05362955, NCT06165614, NCT05413811) and future studies on topical therapies for cervical precancer in LMICs, studies on their acceptability and barriers to uptake among both women and their male partners in LMICs are needed. The objective of this study was to assess how African women receiving cervical cancer screening and precancer treatment perceive the use of topical therapies for cervical precancer treatment and their potential acceptability of such therapies were they to be available.

## Methods

### Study design and approach

This study is part of a larger project exploring the acceptability of topical therapies for the treatment of HPV and cervical precancer, which included in-depth interviews and focus groups with women undergoing cervical cancer screening and male partners in Kenya, in eastern Africa. Results of a qualitative analysis of men’s perspectives have been reported elsewhere [23]. This current analysis encompasses focus group discussions with female participants.[26] We used a constructivist paradigm to gather perspectives of women introduced to the idea of a novel treatment method for HPV or cervical precancer. Constructivism suggests that knowledge is constructed through individual perceptions, experiences, and social contexts [27]. We hypothesized that acceptability of topical therapies is based on women’s experiences (e.g., prior treatment experiences, knowledge of other women’s experiences) and their social contexts (e.g., relationships with sexual partners).

We used focus group discussions (FGDs) to gather the breadth and depth of experiences from groups of women. A predetermined sample size of five focus groups was selected based on evidence indicating that most themes can be captured within a range of three to six focus groups [28]. Since the topical treatment being proposed is innovative within this study’s context, we conducted an analysis of the data using qualitative description, which is highly suitable for enhancing comprehension in a field with limited knowledge [29]. As this method remains focused on the data itself and involves minimal interpretation, qualitative description effectively facilitated our objective of providing a clear and direct account of the participants’ perceptions, thoughts, and experiences.

### Research Team

The principal investigator (CM), a Kenyan-born Obstetrician/Gynecologist with 10 years of experience, graduate students in medicine, social work, and public health (AGK, GZ, SKG), and a senior qualitative investigator with 20 years of experience in qualitative methods and health services research (RMF) comprised the research team. The focus groups were facilitated and transcribed by two qualitative research assistants from the local community.

### Sampling, recruitment, and data collection

We used purposive sampling and a stepped recruitment process to recruit FGD participants, as described previously[23] [30]. Women age 25 to 65 years undergoing cervical cancer screening or precancer treatment in public clinics in western Kenya between November 2022 and April 2023 were included in the study. Emphasis was placed on recruiting women with a history of positive screening results or prior precancer treatment.

Participants were recruited from HIV clinics as well as clinics serving the general population. Most women had undergone cervical cancer screening using HPV self-collection, which was available at most clinics at the time of recruitment. Per the WHO guidelines, women who screened positive were offered treatment with thermal ablation or referred for excision if not eligible for ablation (6). Using focus group discussions (FGDs), we explored the women’s perceptions and hypothetical acceptability of using proposed topical, self-administered therapies for treatment of HPV or cervical precancer, should such therapies become available for public use.

The FGDs were conducted by two female moderators from the same community as the research participants (EA, JO). The moderators had training in qualitative research, prior experience conducting focus group discussions, familiarity with the local context, and fluency in the local languages. FGDs were held at facilities near the recruiting clinics and conducted in the two most spoken local languages (*Swahili* and *Dholuo*). Discussions were guided by several domains of inquiry: 1) baseline knowledge of HPV and cervical cancer screening and prevention, 2) the primary treatment experience and perceived efficacy of treatment, 3) acceptability of self-administered topical therapies as primary or adjuvant treatment to current therapies, 4) self-perceived barriers to use of topical therapies, and 5) perceived barriers or facilitators of male partner’s support for the use of topical therapies as adjuvant treatment. Moderators used standardized language to explain cervical cancer screening and prevention and the potential option of topical self- or provider-administered therapies for precancer treatment. Briefly, participants were introduced to two topical therapies for which data are available, 5-FU and Artesunate, including details on their frequency of use (5-FU once every other week for eight applications, Artesunate daily for five days for three cycles), abstinence requirements (two to three days of abstinence after each 5-FU application and none for Artesunate). Participants were told that tampon use overnight was recommended following application of the topical, and tampons were available for illustration using a pelvic model for those who had never used one. Each FGD included 5-8 participants and lasted approximately 90 minutes. All FGDs were audio recorded, and recordings were transcribed verbatim, translated to English, and crosschecked to confirm accuracy.[31]

### Data Analysis

A codebook was created a priori based on the focus group guide. Two coders (GZ, SKG) read and coded two of the five FGDs to test the code application and gain a sense of additional topics covered in the group discussions, adding emergent codes (e.g., informational needs, interactions with health service providers) to a final codebook. All FGD transcripts were coded using the final codebook. To ensure agreement between coders, a random sample of transcripts was chosen, and the codes were compared for concurrence. Any inconsistencies were addressed through discussion and mutual agreement, and any modifications made were recorded in the codebook. The team reviewed and summarized code reports and explored the data for patterns and themes. Content analysis and thematic development were supported using NVIVO Version 13. Although the focus group discussions covered multiple topics, this analysis focuses on three primary topics: 1) participants’ knowledge and awareness of cervical cancer; 2) treatment preferences and comfort with topical therapy; and 3) perceived acceptability of topical therapy for cervical precancer treatment (Figure 1).

**Figure 1.**
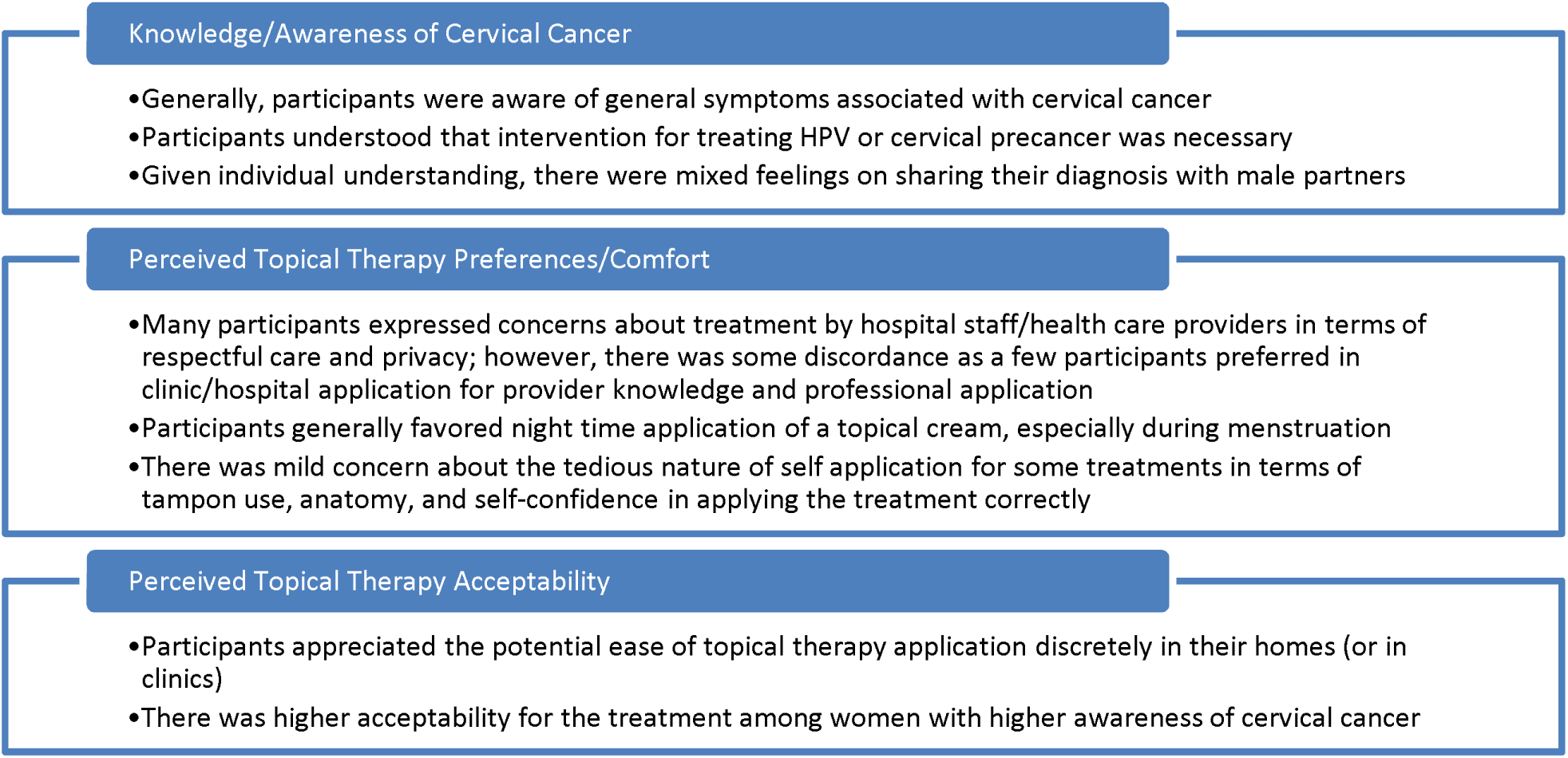
Summary of themes regarding women’s perceptions of topical, self-administered therapies for cervical precancer treatment

## Results

A total of 29 women participated in five FGDs. The mean age was 35.4 years (SD 6.5). The majority, 25 (83.3%), had a history of prior precancer treatment, including during the visit they were recruited into the study. Analysis of the FGDs identified 15 themes related to the potential use of self-administered topical therapies for cervical precancer treatment in the study population, summarized in Figure 2.

**Figure 2.**
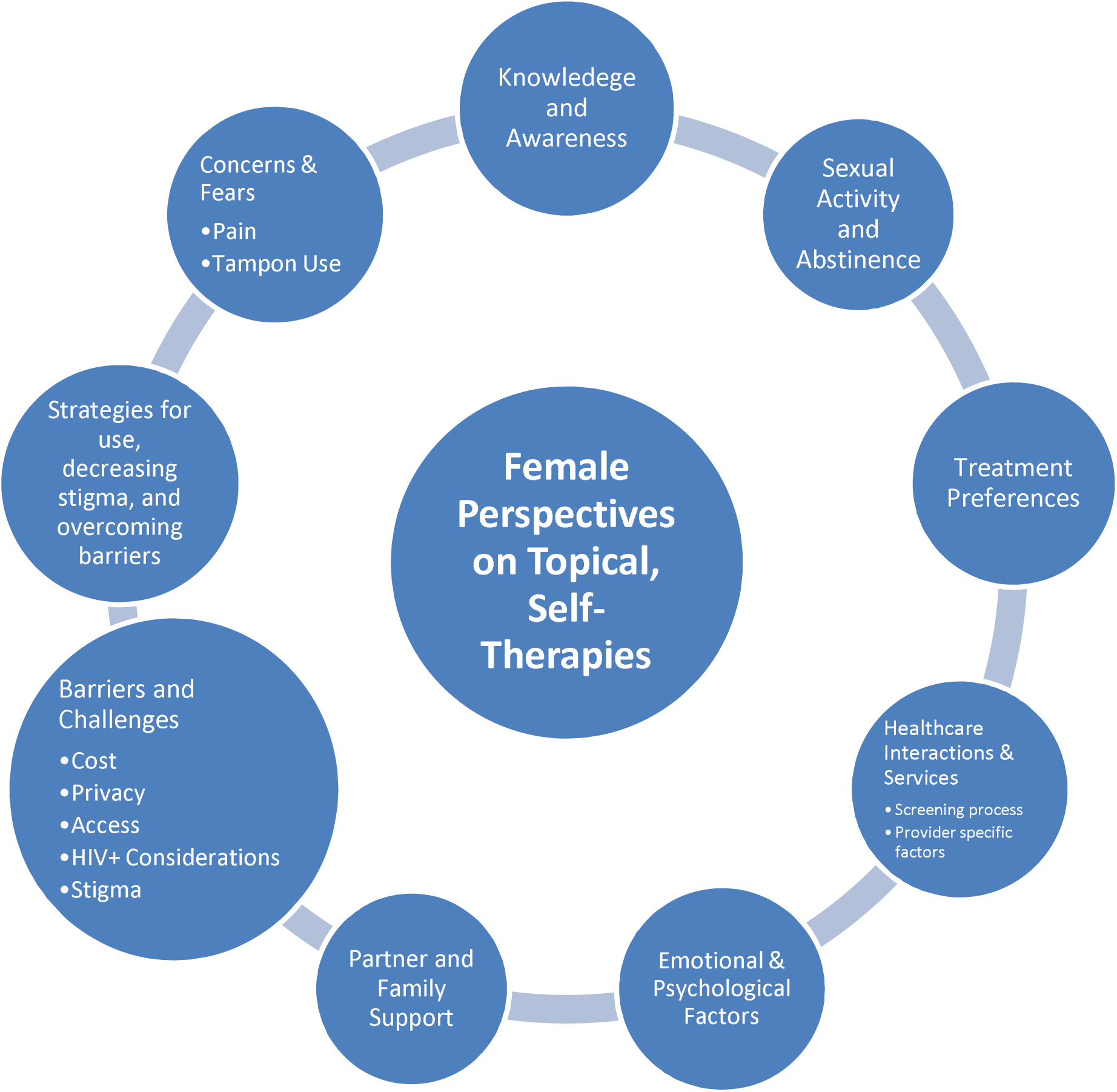
Key findings on women’s general perceptions of topical therapies

### Participant Experiences with Cervical Cancer Screening and Precancer Treatment

FGD participants shared their experiences of learning their HPV or cervical precancer diagnosis following screening. Many mentioned having symptoms of pelvic pain or bleeding during intercourse and wanting to see a doctor for screening and treatment to learn more.

*“I was suspecting something was wrong because I had some pelvic pain and also some spots whenever I had sex, I would have some blood spotting. And I used to hear that those are some of the symptoms suggestive of cervical cancer.”* -R4, FGD1

Others underwent cervical cancer screening after being advised to do so by clinic staff. One woman shared on why she had screening.

*“I cannot refuse because each person just wants good health. They tested and told me that I would be called by somebody after some time.”* -R6, FGD4

While there was acknowledgment and awareness of the symptoms of cervical cancer, awareness of HPV was less common. Following notification of a positive HPV test, some participants noted that they were initially not aware of the difference between HPV and cervical precancer or cancer, and they often needed to seek more information to understand the differences.

*“When I was told that I was HPV positive from the screening test results, I was very afraid from that day and all I could do was to GOOGLE about it and learn as much as I could. But what calmed me down was that when I was being treated for the HPV, I was told that having HPV doesn’t mean that I have cancer. It can be treated early before it progresses to cancer.”*

### - R7, FGD3

Generally, participants shared that knowledge and awareness led to greater acceptance of the recommended treatment, particularly as it relates to the difference between receiving an HPV or cervical cancer diagnosis.

*“I was told that having HPV doesn’t mean I have cancer and so when I went through treatment, I didn’t feel much pain except for the day of treatment, but for a few minutes then I was told to abstain for 6 weeks for the cervix to heal. Then I followed that, and I feel better now.”* - R7, FGD3

Some women recounted their clinic experiences, noting how they felt when they had nice providers compared to others who had previously scared them in some way, an important factor in accepting the news of their screening and the precancer treatment they were prescribed:

*“My test results for my last precancer test are out, and they are positive, and I needed to come for more information and treatment…[the staff] lady who called talked to me…didn’t scare me she talked nicely to me then I came [to the hospital]. I was treated and I was told the discharge will be there for one week, 10 days. But what I felt when I was being treated, I was counseled first, I felt some cramps for some minutes, and I even screamed a little there.”*

### -R2, FGD3

The focus groups highlighted how participants’ experiences varied in learning about their screening results and becoming aware of the treatment options available. The discussions consistently showed that participants’ understanding of HPV and cervical cancer—from prevention through screening to treatment—played a crucial role in overcoming stigma and pursuing treatment after their diagnosis.

### Experiences with Ablative or Excisional Precancer Treatment and Perceived Advantages of Topical Therapies

During the FGDs, participants shared their experiences with traditional precancer treatments (thermal ablation, cryotherapy, excision) and were introduced to intravaginal topical therapies (creams or suppositories) currently being studied that can be self- or provider-administered. Participant’s views on these topical therapies, their potential integration into their lives, and comparisons with traditional treatments were explored.

Many FGD participants showed a greater preference for topical treatments over traditional precancer treatments, which many had undergone, citing topical therapies perceived fewer side effects, especially pain, compared to treatments they had received.

*"What I feared was the LEEP [Loop electrosurgical excision procedure, a surgical precancer treatment method] and secondly the chemoablation [thermal ablation]. A friend who came from treatment would tell us that it is painful, and even the doctor told us that there will be pain during the procedure, especially during heat application. And for sure, [thermal ablation] was painful just as labor pains."* R1, FGD5

*“[With ablation,] during treatment, they were removing certain things [like] cotton wool and in addition to that my sister had also said to me that there is a chemical they will spray, which they did and I felt abdominal pain. I felt pain during treatment, but I just persevered for the sake of treatment so that I get well.”* - R4, FGD4

Many were happy to hear of the potential for a self-treatment option that could be done in their own home, which they felt could better fit into their daily lives, offered more privacy and less discomfort compared to provider-administered treatments:

*“I can prefer cream because that other thermos [thermal ablation] treatment or cryo [cryotherapy], they use strange objects in the cervix and that brings tension and discomfort because the objects going into the cervix makes you tensed and then again that type of treatment [cryotherapy] doesn’t involve one person, you find that three or two people want to deal with your cervix and this brings some discomfort. But this one you are alone with your husband whom you are used to, there is no fear.”* R7, FGD1

“*I can choose [the] cream because it has some level of confidentiality, you know women don’t like it when someone is looking at her private part. So, some people can fail to go back to the hospital because they don’t want the doctor to look at their private part and with [the] cream you apply it yourself and you are the only one who know how your private part looks like [laughter] so I can choose this one”* – R4, FGD1

*“The [treatment at] the hospital that you go to be checked by the doctor, you must at least be seen by a person when on your way there and there is no privacy there. This cream is private,”* -R4, FGD1

Others cited the convenience and accessibility of a topical therapy that can be self-administered at home as an advantage, compared to the time and costs associated with visiting clinics for provider- administered treatments.

“[the] *cream is good due to lack of transportation all the time when going for other treatment methods. Again, I don’t have to make a queue in the hospital waiting to be treated because once I get the cream, I will be applying it by myself at home.”* – R3, FGD4

Some also noted a sense of increased autonomy or empowerment with use of self-administered topical therapies, which some felt would support compliance.

*“If I can be given this cream to take home, nothing can bar me from using [the] cream, I am just being empowered and I use it accordingly.”* – R7, FGD2

“*I think that cream is very good. You cannot fear your own body and therefore you will insert it very well because you want to get cured fully.”* – R5, FGD4

Across all focus groups, the idea of a topical treatment applied at night was embraced, citing convenience as it meant that the day’s activities would be over.

“*I feel [it] is better at night because you are just resting, I don’t like using it daytime because I will be walking maybe the medicine can flow [out], and that is not good*.” - R5, FGD2

“*Applying it at night is very good because it is a time that I am retiring to bed. Secondly during the day, I will pass urine a lot but in the night once I have put it [applied the cream? Used the tampon] I know it is [in place?] until morning. Since I am the one who chooses my best time that suits me…even if I have children, they would have slept by then, even if I have a husband we will be just the two of us. I am the one to choose one that suits me.”* – R4, FGD3

Although most focus group discussion participants favored self-administered treatments, some expressed a preference for provider-administered thermal ablation, emphasizing their comfort in trusting a doctor to accurately apply the treatment to the correct area of the cervix.

“*The reason I may only like the one I was done for [thermal ablation], than this [self- administered cream], that one was done by the doctor, when she doesn’t see well, she cleans and confirms, if not well done she does it again until it reaches where she wants, but using cream, you are in the dark, you don’t know whether you have placed it well or not.”* – R6, FGD2

Similarly, participants who favored having a topical treatment applied in the clinic by a healthcare professional compared to self-application expressed confidence in a doctor’s ability to administer it more effectively than they could themselves, particularly if they encountered side effects during the application, which may cause them to hesitate with self-administration.

“*Sometimes you can decide to try it [topical therapy] a little bit and see how it is, if you find it itching you might stop using it. And you see with the doctor he will just go ahead and apply it, and once he applies it is done.”* – R3, FGD3

Generally, the majority of focus group participants were open to using a self-administered treatment if it were accessible. Crucially, they noted that self-administered treatments at home could shield them from adverse interactions with healthcare providers in clinics, such as being shouted at, which some had experienced while seeking treatment for precancer.

*“Women can be free to apply it [topical therapies], they have their own time without worry of meeting a doctor, maybe one who shouted at her last time…What I felt when I was being treated [with non-topical treatment], I was counseled first, I felt some cramps for some minutes, and I even screamed a little there. The health providers touched my pelvic [area] and asked me to chill.”- R*2, FGD3

### Participant’s Preferences Between Two Proposed Topical Therapies

In the FGDs, participants were introduced to two potential self-administered topical therapies for cervical precancer treatment: topical 5FU cream and Artesunate suppositories. The differences between the two therapies were described, including treatment length (5FU is used once every two weeks for 8 applications over 16 weeks, while Artesunate is used nightly for 5 days, followed by a week off, repeated for 3 cycles over six weeks), and abstinence requirements (abstinence is required for 2 days after 5FU use, while abstinence is not required with Artesunate use). Participants were then asked which of the two potential therapies they would prefer, if they needed to use it, based on these described characteristics.

Those who preferred the 5FU treatment did so because of the perceived ease of the application regimen – once every two weeks for 16 weeks, compared to daily use for Artesunate.

*“What makes it [5FU] better than the other one [Artesunate] for me is that maybe you have traveled, so you know the one for once every two weeks, even if you apply it, even if you are on a journey, it does not worry you.”* – R3, FGD2

*“I think 5 FU is good especially for those are not held up in their minds, there those who are busy all the time and since the 5 FU is not complicated, for the AS [Artesunate], it is complicated, you can forget the days, again with the menses disruption, it is not the best. I think 5FU is the best option.”* – R1, FGD4

*“[The 5FU] will suit me because I don’t have to put it all the time.” -* R2, FGD3

“*…[I] am comfortable with it once a week, for the daily one you might have some occasions like funeral and finding a place for you for application may not be easy.”* – R7, FGD5

Others preferred Artesunate because of its use over a significantly shorter duration – only 6 weeks compared to 16 weeks for 5FU- and the possibility that condom use may not be required with its use

*“Though the [Artesunate] is a bit tedious, you are using the medication daily, but it is a shorter period of time then it doesn’t have a lot of restrictions.”* R6, FGD1

“*The treatment that I would prefer is [Artesunate], the one where you treat for 5 days then the following week you rest, then you also don’t use a condom and it is a shorter period of treatment than the one that goes for 16 weeks. Though the 16 weeks also have weeks when you are skipping but it is a long period then it has condom use for the whole treatment period. So, for me because condom will cause conflicts in my house, I would settle for [Artesunate].”* – R7, FGD1

Participants noted that their treatment preferences were influenced by their perceptions of their male partner’s opinions of such therapies, including the requirements for condom use. Many cited that abstinence for long periods of time could be a source of conflict with their male partner’s preferences. This was cited as a reason why therapies like Artesunate, which may not require condom use or abstinence, may be preferable over 5FU which requires both for certain periods:

“*I like where there is peace…but maybe the way the husband as we were saying, they might not understand the abstinence part and even this condom use, they usually say that they cannot use a condom with their partners, they feel like if you insist then there is something and this alone can cause conflicts. So, I prefer [Artesunate] even if I am applying for 5 days in peace, it is better because I know he is going to support me, and the medication will work well than the one where you are fighting. And you know there are some that might even end up breaking the rules, so peace is good*.” – R6, FGD1

“*For me condoms can cause conflict, most men don’t like using a condom and there are those who have never used a condom in their life.”* – R7, FGD1

Participants who did not favor Artesunate pointed out the inconvenience, especially the burden of applying it every day. Furthermore, those with irregular menstrual cycles noted that 5FU was more manageable due to its biweekly application schedule, which is simpler to follow than Artesunate’s daily regimen, which can be interrupted by irregular periods.

### Considerations for Women Living with both HIV (WLWH) diagnosed with HPV

In the FGDs, participants who were living with HIV (WLWH) who had also tested positive for HPV or cervical precancer noted feeling an increased burden. Many expressed fears about the impact of the dual diagnosis on their children or other family members, as well as the challenges of managing multiple medications when treating cervical precancer alongside HIV infection.

*“It also bothered me, and it stressed me out following that I am also on HIV medication, I felt very bad because I also infected my baby [with HIV]. I have been taking HIV medication from 2009 up to now. So, when I imagined getting another terminal illness, I felt sad.”* – R6, FGD3

*“And if I consider that I had [pre]cancer and with HIV, it was double burden. The fact that cancer can worsen and kill you I get very bad. And if I consider that I had [pre]cancer and with HIV, it was [a] double burden and so, I decided to clear with [pre]cancer which is curable.”* - R7, FGD5

One participant believed that topical treatments for cervical precancer would be insufficient due to their concurrent HIV and HPV diagnoses. They harbored doubts about the effectiveness of such treatments when dealing with both conditions simultaneously.

*“According to me I feel the [topical] treatment is not 100% for those who have HIV, because of our low immunity, our system is weak. So even if we are treated, we can still just get [cancer].”* – R7, FGD3

Other participants likened the use of self-administered topical therapies among HIV-positive women to the same way WLWH are prescribed antiretroviral therapies (ARVs), which they use at home to treat their HIV disease. The participants drew parallels between their consistent use of ARVs at home and their potential to similarly apply self-administered topical therapies in the same settings.

*“Those who are HIV+ should go for [the topical] cream because, they go to the hospital for ARVs refill, they should take cream and use it at home just the same way they take ARVs and adhere to its use at home.”* R2, FGD4

“*ARVs is for HIV and cream is for HPV or precancer, so you just take your medication and also apply your cream because they treat different things.” –* R7, FGD3

Generally, participants ultimately felt that the time required to apply the topical therapies was shorter in duration in the home setting versus returning to the clinic to be treated, which greatly influenced their perceived acceptability and desire to use topical therapies. Regardless of HIV-seropositive status, the participants noted that if they had the knowledge and the ability to apply the cream at home, they would be willing to do this for the betterment of their health.

## Discussion

In this qualitative study evaluating Kenyan women’s perceptions of topical self- or provider-administered therapies for cervical precancer treatment, we find that participants, many of whom had undergone traditional cervical precancer treatment, were highly receptive to topical therapies. We found that many participants had fears following a diagnosis of HPV or cervical precancer, which they had to overcome in order to undergo ablation or excisional treatment procedures. When introduced to topical therapies as a potential alternative to available precancer treatments, participants strongly favored topical therapies, citing reduced pain, improved accessibility, and privacy, compared to the currently available provider-administered precancer treatment methods that many had undergone. Most study participants expressed a strong preference for self-administration of topical therapies, with many citing the lack of privacy associated with provider-administered treatments as a barrier that those who had received precancer treatment had to overcome and that often discourages other women from seeking treatment. Participants ’ preferences varied when given an option between two potential topical therapies with different characteristics and requirements for use. Some favored 5FU, applied every two weeks, despite its conditions for abstinence following use and consistent condom use. Meanwhile, others favored Artesunate, which requires more frequent applications but may have less stringent restrictions around abstinence and condom use. Despite only having had a brief education session, participants showed high levels of awareness and body autonomy in the discussions by displaying keen insights into potential different trade-offs associated with the two topical therapies discussed, including the impact of irregular menstrual cycles on the ability to adhere to a daily topical. Given the higher incidence of cervical precancer in women living with HIV, it is noteworthy to highlight that HIV-positive participants in our study indicated concerns about managing their HIV disease alongside a diagnosis of HPV or cervical precancer. However, most were confident about their ability to use a self-administered topical treatment for cervical precancer, drawing on their experience with daily use of oral antiretroviral therapy to manage HIV infection.

To our knowledge, this is the first qualitative study to explore African women’s perceptions and perceived acceptability of self- or provider-administered topical therapies for cervical precancer treatment. In this study of urban and peri-urban Kenyan women who had undergone cervical cancer screening and a majority of whom had undergone ablation or excisional precancer treatment, many expressed conflicting emotions about their treatment, explicitly highlighting the challenges they had to overcome in terms of access, pain and lack of privacy often pointing to pain and privacy issues when receiving provider-administered treatments. Most showed a preference for topical therapies, if available, believing they would alleviate these challenges associated with conventional treatment methods that often deter other women from pursuing precancer treatment. The acceptability of thermal ablation, the most widely available precancer treatment method in LMICs that was approved by the WHO in 2019, has been demonstrated in a few studies[28], [29]. Thermal ablation, which involves the application of a heated probe to the cervix to destroy precancerous tissue, is performed without local anesthesia to the cervix[6]. Studies in LMICs report that while 83.9% - 90% report no or mild pain with thermal ablation, 2.5% - 16.1% report high or moderate pain with the procedure [28], [29]. In our qualitative findings, some participants described thermal ablation as “*too painful*” or “*painful just as labor pains.*” Another participant noted the need to encourage women that the procedure is “*not very painful*” and should not deter them from presenting for treatment, as the pain perception is thought to keep women away from presenting for treatment. Studies on whether certain women undergoing thermal ablation may require pretreatment analgesia are needed, alongside considerations of the feasibility of providing of doing this. If topical therapies for cervical precancer can be shown to be equally effective as ablative or excisional procedures in low- and middle-income countries (LMICs), they could potentially alleviate the pain-related concerns associated with ablation or excision.

Our findings of participants noting challenges with treatment access and privacy concerns associated with provider-administered, facility-based treatments have been demonstrated in several LMIC studies. Facility-based precancer treatment access challenges in LMICs include lack of functional equipment or supplies[10], [13], lack of trained providers [7], [10], [13]long distance required to access treatment facilities [11], [30]. These factors significantly contribute to the existing precancer treatment gaps. In a study from rural Kenya, up to 40-50% of women who screened positive and were referred to a central facility did not make their follow-up appointment [11]. Similarly, in a qualitative study from Malawi, women with abnormal cervical cancer screening results cited lack of transportation to referral facilities and high associated costs as major reasons for not presenting for treatment [30]. This is reflected in our study, where women emphasized the convenience of self-administered topical therapies that can be used at home, highlighted ease of access to topical self-administered therapies used at home, compared to facility-based treatments, which are associated with high transport costs and long waiting times at the facilities as a reason they would favor topical treatments. Similarly, our findings of increased privacy as a reason women prefer self-administered therapies to conventional treatments have been highlighted in prior studies, which found that fear of a violation of privacy associated with pelvic exams [31], [32], [33], [34], and especially when performed by a male provider [33], [34], [36], are barriers to screening and precancer treatment in sub-Saharan Africa. As noted by a study participant, during her ablation procedure, “*two or three people want to deal with your cervix, and this brings discomfort*,” stating that with a self-administered treatment, “*you are alone with your husband whom you are used to, there is no fear*." The use of self-administered topical therapies, which women can apply in the comfort of their own homes, can be a scalable way to address both the access challenges and privacy concerns of African women.

Self-administered therapies can also promote women’s autonomy and sense of agency, as highlighted by our study participants, who stated that they anticipated "*feel[ing]empowered*” and would use it correctly, as “*you cannot fear your own body.*" The use of self-administered precancer treatment, if backed by feasibility and efficacy studies in LMICs, also aligns with a recent guideline from the World Health Organization that advocates for self-care interventions. As stated in the guideline, these interventions have the capacity to “increase choice and autonomy,” address the global shortage of healthcare workers, and bring us closer to achieving universal health when made “accessible, acceptable and affordable[37].” While no studies have evaluated the acceptability of self-administered topical cervical precancer treatment in LMICs, several studies in this setting have demonstrated high acceptability of self-care interventions, including HIV self-testing [38] and the use of vaginal or rectal microbicides for HIV prevention[39], [40]. Similarly, in a study on the acceptability of rectal microbicide for HIV prevention among men who have sex with men in Thailand, ease of use, privacy, and comfort of use at home were facilitators of uptake [41], drawing similarities to our findings.

This study has several strengths, such as the inclusion of women who have had cervical cancer screening, as well as a deliberate oversampling of women with a history of precancer treatment. This approach ensures that the study represents the demographic that is most likely to benefit from topical therapies, hence whose perceptions are important in understanding acceptability. Similarly, the use of focus groups in the qualitative design facilitated in-depth discussion among study participants who shared similar experiences. This enabled the identification of multiple themes that impact the acceptability of this intervention to inform feasibility studies. The study’s inclusion of women living with HIV is a significant strength due to their higher risk of cervical cancer and current unmet need for accessible precancer treatment. A limitation of this study is that participants expressed theoretical acceptance of the intervention but did not actually use the topical therapies. Therefore, their views might change with actual use, an aspect future studies should explore. Another limitation was the limited time for focus groups; more time could have offered insights into household dynamics like decision-making and empowerment, potentially affecting women’s perceptions of the therapies.

## Conclusion

Our findings from Kenya indicate that women find these therapies acceptable and that they have the potential to address significant challenges like access, privacy, and cost that hinder precancer treatment uptake in these regions. These results support ongoing feasibility studies and call for efficacy studies in this population to inform whether these treatments can be made available to women.

## Data Availability

Data are available upon reasonable request.

## List of Abbreviations

5FU: Fluorouracil
FGD: Focus Group Discussions
HIV: Human immunodeficiency virus
HPV: Human papillomavirus
LEEP: Loop electrosurgical excision procedure
LMIC: low-and middle-income countries
SD: standard deviation
WHO: World Health Organization
WLWH: women living with HIV

## Declarations

### Ethics approval and consent to participate

#### Ethical Considerations

The study was approved by the ethics review boards at Maseno University School of Medicine in Kenya and the University of North Carolina Chapel Hill in the U.S.A. All participants provided written informed consent prior to study participation.

#### Consent for publication

Not applicable

### Availability of data and materials

Data are available upon reasonable request. For inquiries or to request access to the data, please contact Chemtai Mungo at.

## Authors’ contributions

CM conceptualized the study and associated clinical trials. EA and JO conducted focus group discussions with study participants . GZ, SKG, RMF, and AGK worked on qualitative methodology, with GZ and SKG conducting data analysis and RF leading methods section in this manuscript. AGK led manuscript writing with CM and RMF. CM, AGK. EA, GZ, SKG, JO, and RMF all read and approved the final version of the manuscript.

## Acknowledgments

The research participants and leadership of Lumumba Sub-County Hospital in Kisumu, Kenya, are acknowledged.

